# Genome-wide characterization of clonal hematopoiesis reveals extensive non-coding putative driver mutations

**DOI:** 10.1101/2025.10.12.25337792

**Authors:** Joshua S. Weinstock, Karen Conneely, Janghee Woo, Marios Arvanitis, Mitchell J. Machiela, Cameron Russell

## Abstract

As humans age, we acquire somatic mutations in our blood, leading to clonal hematopoiesis (CH). Despite the prevalence of CH in aged individuals, recent searches for selective sweeps in single-cell derived colonies have revealed that most clones have expanded without a known driver mutation. This extensive, unexplained CH motivated our search for novel driver mutations across ∼490K blood whole genome sequences from the UK Biobank. We searched across variants with a minor allele count of at least 10 (∼147M variants) to discover alleles that are enriched in aged individuals. We identified 45 variants including known CH driver genes (e.g., *DNMT3A*, *ASXL1, SF3B1*) and 35 novel variants.

Among the novel age-associated variants, we identified 72 carriers of a somatic mutation in the *TERT* promoter. Although previously reported in pan-cancer analyses, *TERT* promoter mutations have typically been excluded from population searches for CH. We also observed a somatic intronic insertion in *UGT2B7* in 1,165 carriers, a cluster of *IGH* point mutations, and centromeric variation. We conducted a phenome-wide association study (PheWAS) among 30 common disease phenotypes to characterize the phenotypic correlates of these mutations, finding 965 links between somatic mutations and common diseases, including 37 protective associations. After estimating the total liability scale variance explained of CH mutations on each common disease, we found that non-canonical CH contributed 28% of the variance explained. We then performed a genome-wide association study of the *IGH* mutations, finding that common germline variation at *GRAMD1B* is strongly associated with *IGH* mutations, and finally a proteome-wide association study to characterize the plasma proteomic correlates of CH. Overall, we characterize CH at both increased breadth and resolution and characterize the entire cascade from upstream germline risk haplotypes to downstream clinical correlates. We release our summary statistics in a publicly accessible portal, somatic.emory.edu.

## Main Text

As humans age, hematopoietic stem cells (HSCs) acquire somatic mutations, some of which result in an expansion of a mutated lineage of cells (“clone”). Recent analyses of hematopoiesis across differently aged human donors have revealed that as we age, hematopoiesis transitions from polyclonality to oligoclonality^1^. This phenomenon is commonly referred to as aging-related clonal hematopoiesis (CH), and has recently been invoked as a novel contributor to disease across several distinct organ systems, including hematologic malignancy^2–7^, cardiovascular disease^8–11^, kidney disease^12^, liver disease^13^, among others. Initial CH discovery efforts sought the characterization of specific ‘driver’ mutations, reporting a class of recurrent point mutations across ∼80 genes^14^, along with genetic lesions altering large stretches of DNA sequence, including mosaic chromosomal alterations^15–19^, loss of the Y chromosome^6,7,20^, and loss of the X chromosome^21^. However, sequencing efforts of single HSC derived colonies^1^ have revealed that the majority of CH occurs in the absence of known driver mutations, indicating the current characterization of the etiology of CH is largely incomplete. Concordant with these observations, evolutionary modeling of the abundance of clonal synonymous mutations^22^ indicates substantial positive selection in HSCs that is unexplained by known genetic lesions. Emerging theories for unexplained clonal expansions include the possibility of epigenetic alterations (‘epimutations’) sufficient to alter HSC fitness^23^, along with the possibility of undiscovered somatic driver mutations.

Initial searches for genetic drivers focused on coding mutations in manually curated lists of genes with links to hematologic malignancy^2,3^. Recent efforts have been defined by the ratio of non-synonymous to synonymous (dN/dS) mutations across all protein coding genes, resulting in the discovery beyond ‘canonical’ CH genes^24^. However, no analysis has searched across the entire genome for drivers; searches defined based on dN/dS exclude non-coding mutations, and are likely biased towards longer genes^25^ which have more opportunities for non-synonymous mutations resulting in differences in statistical power.

Although searches for non-coding drivers have occurred in pan-cancer cohorts^26^, few analyses have performed in the biobank CH context^27^.

In this work, we performed an unbiased search for drivers across ∼147M coding and non-coding variants detected in ∼490K blood whole genomes sequenced by the UK Biobank^28^ (UKB). We defined drivers as those variants that rapidly increased in frequency during aging, which previous evolutionary models have demonstrated is highly unlikely to occur due to stochastic drift^29^. We discovered 35 novel drivers, including an intronic insertion in *UGT2B7*, a mutation in *TERT* promoter, and non-coding mutations in the Immunoglobulin Heavy Chain (IGH). We then comprehensively mapped the common disease correlates of these mutations, performing a phenome-wide association study (PheWAS) among 30 common aging-related disease phenotypes, finding 881 links. We then quantified the relative fitness estimates of these mutations using the passenger approximated clonal expansion rate^30^ (PACER). Finally, to discover the inherited determinants of these mutations, we performed GWAS of the *IG* mutations, finding that common genetic variation at *GRAMD1B* was associated. Overall, we perform the first genome-wide search for age-related CH driver point mutations and their germline and disease correlates across ∼490K genomes. An interactive web portal is now available at somatic.emory.edu.

## Genome-wide search for novel drivers at single-variant resolution

Driver mutations were defined as somatic variants with alleles strongly enriched in older individuals, indicating selective clonal sweeps during aging. We included all SNP and indel variants present in at least 10 carriers (n = 147M variants) among the ∼490K UKB individuals with whole genome sequences and performed a genome-wide association study of the age at recruitment using a linear mixed model, resulting in 68 mutations (**Figure 1**) where the alternate allele was directly or inversely associated with age at genome-wide significance (5 x 10^-8^). Among the 68 age-associated variants, 23 variants were likely of germline origin, as they were substantially more frequent and had smaller effect sizes than the 45 other age-associated variants. Among these 23 likely germline variants, some were linked to genes previously discovered in GWAS of longevity phenotypes^31^, including variants linked to *APOE* (chr19_44909967_TGG_T, pvalue = 3.0 x 10^-8^ Table S1) and *APOC1* (chr19_44919689_A_G, pvalue = 3.3 x 10^-8^, Table S1). Among the 45 other likely somatic variants, we observed several canonical CH genes, including *DNMT3A, ASLX1, GNB1, IDH2, SF3B1,* along with a gene linked to “lymphoid CH”^32^ (*IGLL5*).

**Figure 1:**
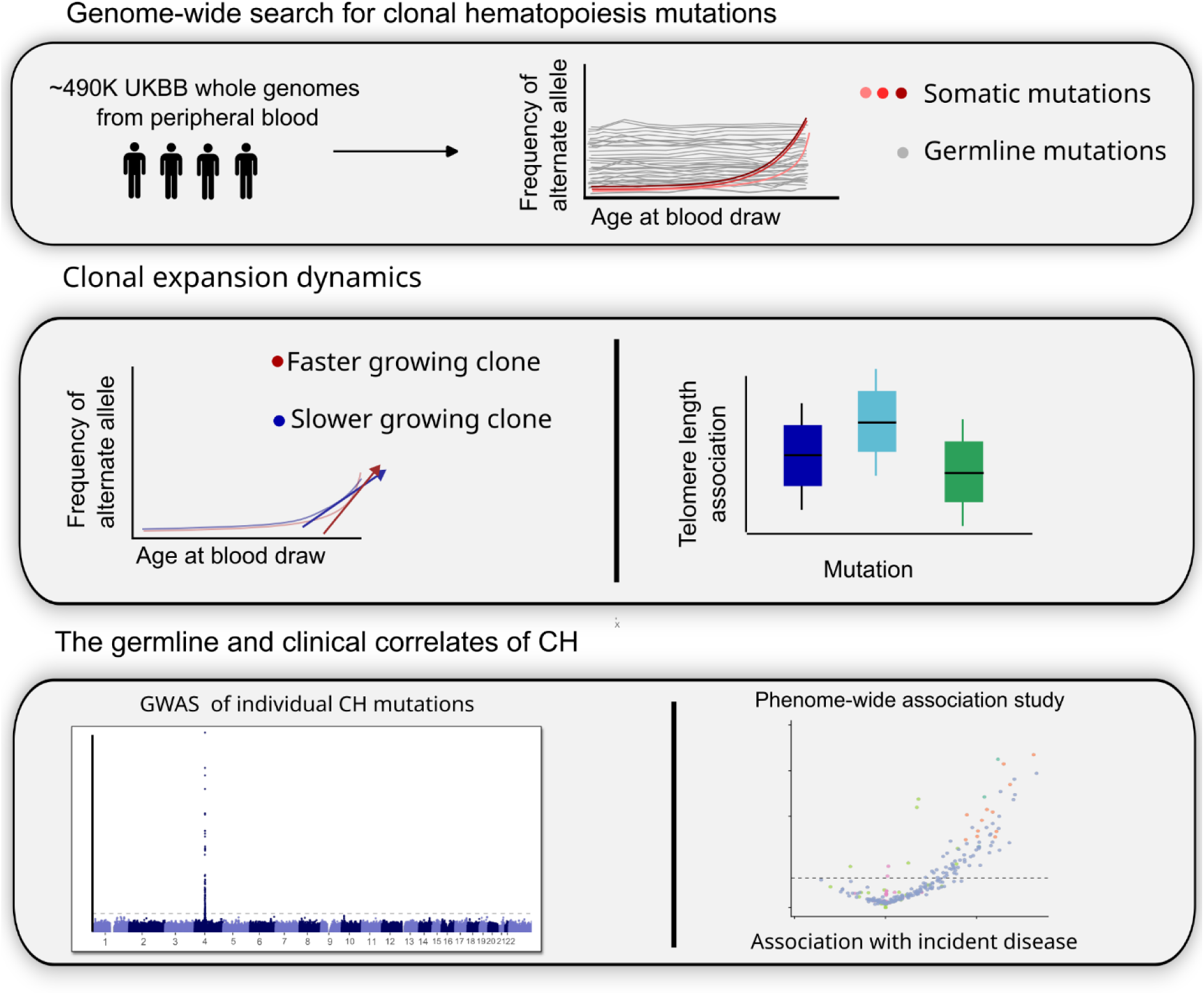
Study overview. **Top**: somatic mutations are called genome-wide by searching for association with age at blood draw. **Middle**: Rates of clonal expansion are estimated using both telomere lengths and the passenger approximated clonal expansion rate (PACER). **Bottom**: GWAS and PheWAS analyses were performed to characterize the germline and phenotypic correlates of clonal hematopoiesis.

As a subset of canonical CH mutations were excluded from the Illumina Dragen output used to define the variant call set, we then performed a secondary targeted calling of known CH drivers using a pileup-based approach (Methods) and included these along with the 45 age-associated variants in all downstream analyses. Among the 45 variants, 43 were more frequent in older individuals, and two were less frequent in older individuals than young (chr2_235126205_A_AG, chr10_133690123_G_T). Out of the 45 variants, 35 are not included among canonical CH driver gene lists^14^, and we refer to them as putative driver mutations.

Among the 35 novel putative driver mutations, we observed an indel in an intron of *UGT2B7* in 1,165 carriers (chr4_69057335_T_TAC), a mutation in the promoter of *TERT*, five *IGH* mutations (four of which are non-coding: chr14_105860408_G_C, chr14_105860409_C_G, chr14_105863341_C_G, chr14_105864020_G_A), and 17 centromeric variants (**Figure 2A-C**, **Supplementary Data 1**). Collectively, these mutations were all rare (frequency < 0.5%) and had large effect sizes (|𝛽| > 0.10 standard deviations in age at recruitment). Of the 35 putative drivers, two are missense (*IGHV2-70D*, *IGLV3-1*), one may affect splicing (chrX_155066068_C_T, *MTCP1*), and the other 32 are non-coding (**Supplementary Figure 1**).

**Figure 2:**
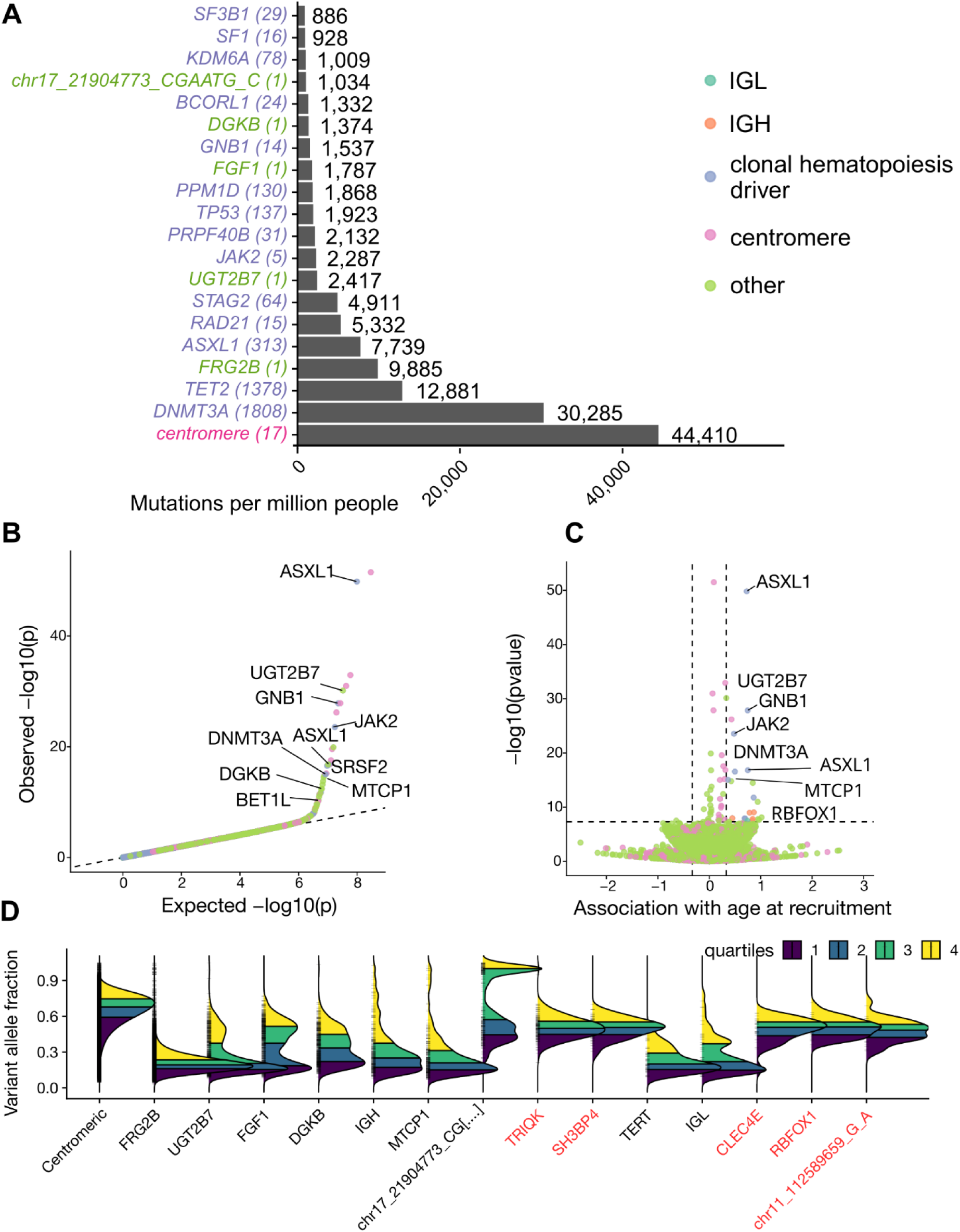
Genome-wide search for clonal hematopoiesis identifies novel putative drivers. ***A.*** The number of individuals with at least one mutation present in the annotated genes per million people. Variants were classified as CH based on a genome-wide search for alleles that were associated with age at blood draw. Canonical CH driver mutations were called with a pileup approach at candidate loci, and other CH mutations are called by the Illumina Dragen variant caller. Genetic ancestry principal components were included as covariates. All variants (n = ∼147M) with at least 10 minor alleles were included, and REGENIE was used to estimate test statistics. ***B.*** quantile-quantile plot from the discovery analysis, indicating limited inflation (*λ*_𝑔𝑐_ = 1.01). ***C.*** A volcano plot displaying the effect sizes in terms of standard deviations of age at blood draw. ***D.*** Distribution of variant allele fractions (mutated reads / depth) are plotted for novel classes of mutations. Hypothesis tests assessing whether the distributions were compatible with germline heterozygous genesis were performed with a likelihood ratio test (**Methods**); mutation groups where the test failed to reject the null are plotted in red.

We then asked whether the novel mutations were merely hitchhiking on clones with driver mosaic chromosomal alterations (mCAs). To assess this hypothesis, we cross referenced the novel putative drivers with mCAs called in the same samples^17^. We observed that the five *IGH* mutations overlapped a relatively frequent mCA (0.25% frequency), suggesting that they may hitchhike on mCA-IGH clones, which is concordant with the presence of a cluster of age-associated point mutations (**Supplementary Figure 2**). In contrast, the *FRG2B, UGT2B7, DGKB,* and *FGF1* linked mutations did not overlap frequent mCAs (**Supplementary Figure 3**).

In contrast to germline variation, where the proportion of reads containing alternate alleles occurs in expectation 50% of the time for heterozygotes in diploid regions, somatic variation is likely to depart from this expectation, as the mutated reads are likely present in only a fraction of sequenced cells (sub-clonal). We used this signature to assess whether these mutations were truly somatic in origin. After performing beta-binomial hypothesis tests (**Methods**), we observed that five of the 35 putative drivers had variant allele fractions (VAFs) compatible with germline genesis (**Figure 2D**, chr8_92974059_GT_G; *TRIQK*, chr2_235126205_A_AG; *SH3BP4*, chr11_112589659_G_A, chr16_6647985_G_A; *RBFOX1*, chr12_8544303_C_CA; *CLEC4E*), suggesting that they could be germline despite their association with age, or might reach these proportions due to allelic imbalances associated with overlapping mCAs. Notably, all centromeric variants had VAFs that were greater than 0.5.

We then sought to validate that our age associated variants are present in other large-scale sequencing cohorts of blood by cross-referencing variant calls derived from 184,878 blood whole genome sequences generated by the NHLBI TOPMed consortium^33^. Importantly, TOPMed uses distinct sequencing and variant calling procedures that differ from the UKB calls, suggesting that presence in both TOPMed and UKB is unlikely to be explained by shared variant calling artifacts. We found that among the 45 age-associated variants, 43 were also present in TOPMed. We observed that alternative allele frequencies were highly concordant among non-centromeric variants (*ρ* = 0.63, *p* = 0.008, **Supplementary Figure 4**). However, we observed that centromeric variants were reported at a much greater frequency in TOPMed than in UKB. We then recalled the age-associated centromeric variants in UKB using an orthogonal pileup based variant calling procedure, finding that coverage of reads at the locus was strongly associated with both age at recruitment and genotype (**Supplementary Figure 5**), suggesting that the centromeric point mutations may tag broader age-associated copy number variation at the chromosome 17 centromere (chr17-CEN).

To assess this hypothesis, we then quantified coverage at the chr17-CEN, which we normalized within sample by estimating the centromere coverage relative to the coverage at *Oct4* (Methods), which was selected due to its intolerance to copy number variation. We similarly estimated normalized coverage estimates for *TET2, DNMT3A, and JAK2*, along with the sex chromosomes. Given the challenges of aligning short-read sequence to centromeric regions, we then performed sex stratified analyses to assess whether chr17-CEN may simply tag incorrectly mapped sequence from sex chromosomes. We observed that chr17-CEN depth is negatively associated with mosaic loss of Y (LOY) in males (beta = −0.008, *p* = 5.3 𝑥 10^−4^), and that it is strongly associated with relative depth at *TET2* (beta = 0.199, *p* < 1 𝑥 10^−300^, **Supplementary Figure 6**), and chr17-CEN coverage is associated with greater age at blood draw (*p* = 3.0 𝑥 10^−15^). Taken together, these observations suggest that centromere coverage may tag a broader genomic instability phenotype, although long-read sequencing technologies may be needed to more carefully evaluate this hypothesis.

Given the challenges in assigning causal genes to non-coding variants, we then applied AlphaGenome^34^, a recently developed sequence-to-function deep learning model, to classify the most likely causal gene in *cis* of the associated variants. We used AlphaGenome to predict the effects of each mutation on the expression of genes within a 1 Mb window of the mutation in hematopoietic multipotent progenitors (Methods, **Supplementary Data 2).** Given that the *TERT* promoter has previously been reported in pan cancer analyses^26^, we selected this region as an exemplar locus, as it contains one of the 35 putative drivers (chr5_1295113_G_A). As anticipated, AlphaGenome predicted that this SNP is likely to upregulate *TERT* in hematopoietic progenitor cells, resulting in increased telomere lengths (**Supplementary Figure 7**). Surprisingly, for many of the variants, AlphaGenome predicted altered regulation of multiple genes. For example, chrX_155066068_C_T is predicted to increase the expression of both *MTCP1* and *CMC4,* and the intronic indel in *UGT2B7* is predicted to both down-regulate *UGT2B7* and up-regulate *UGT2B11*. Notably, *MTCP1* is a member of the *TCL1* family, and is paralogous to *TCL1A*, which we previously discovered as a potent regulator of clonal expansion in CH^30^.

Finally, we asked whether gene-based analyses would result in additional discovery. We performed two gene-based tests; first, we used the aggregated Cauchy association test (ACAT) ^35^ to meta-analyze the single-variant results within each protein gene, and we limited variants to those that were predicted to have moderate deleteriousness by AlphaMissense^36^ (Methods). Second, we used ACAT on all rare variants (MAF ≤ 0.5%) within each protein coding gene. After adjustment for exome-wide significance (2.5 𝑥 10^−6^), we observed that gene-based analyses did not increase discovery over the single-variant analyses.

## Clonal dynamics of driver mutations

Several recent reports have demonstrated that larger clones are much more likely to associate with disease than smaller clones^8,11,37^, underscoring the importance of estimating the rate of clonal expansion. Using a method we previously developed to estimate the relative fitness of driver genes based on quantifying the enrichment of hitchhiking passenger mutations, an approach referred to as the Passenger Approximated Clonal Expansion Rate^30^ (PACER), we then estimated the fitness of the novel putative driver genes referent to *DNMT3A* R882H (chr2-25234373-C-T, **Methods**). Consistent with previous reports, mutations in splicing factors (*SF3B1, SRSF2, PRPF8)* and *JAK2* had larger PACER estimates than *DNMT3A* R882H (**Figure 3A**). Among the putative novel drivers, the *TERT* promoter mutation is estimated to have similar fitness to splicing factors and *IDH2,* and greater fitness than *JAK2.* To our surprise, the *IGLL5* and *IGH* mutations had larger fitness estimates than even the splicing factors.

**Figure 3:**
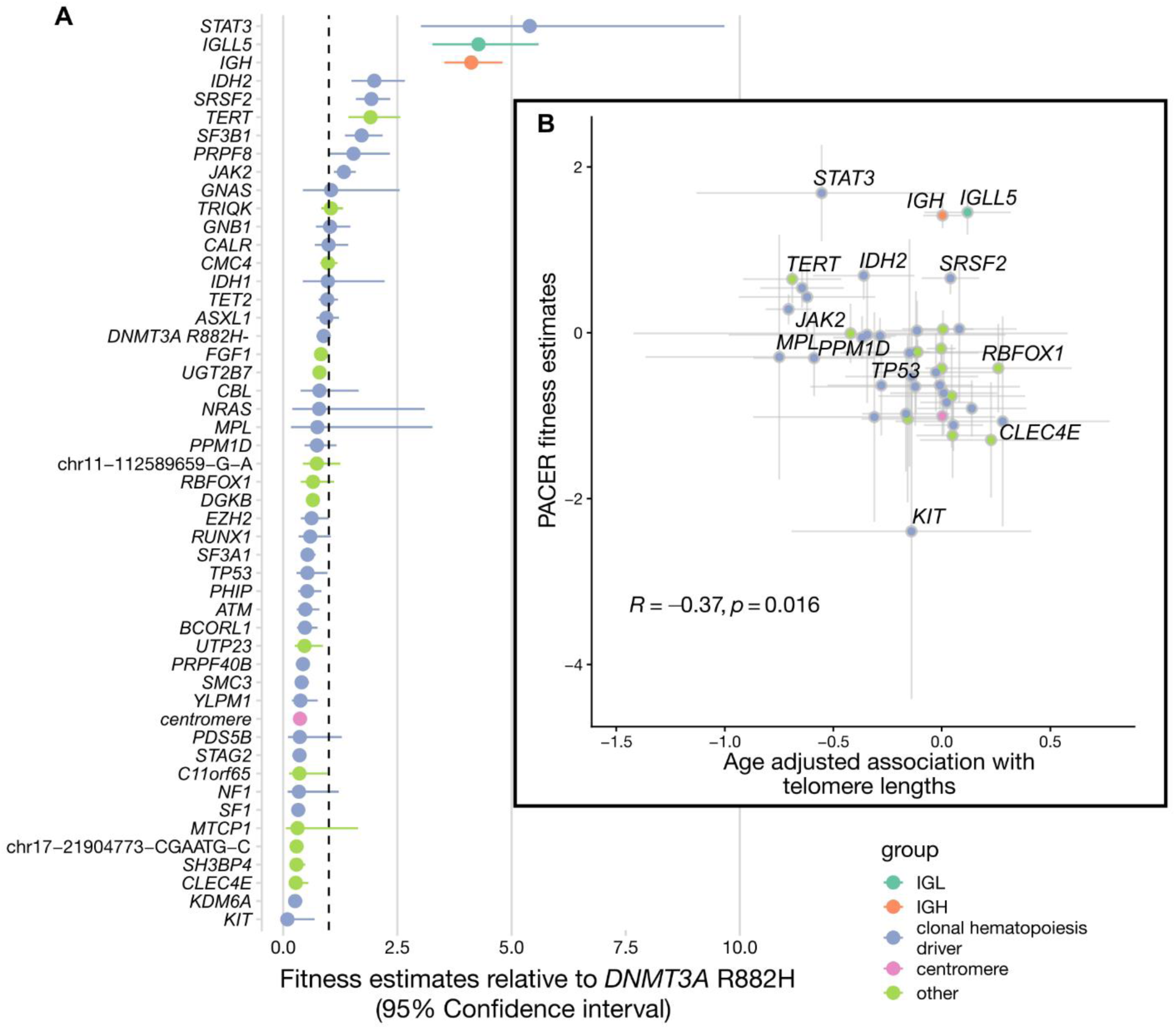
Fitness spectrum of clonal hematopoiesis. ***A.*** PACER estimates of relative driver gene fitness, with DNMT3A R882H as the reference. Excess passenger mutations are estimated using negative binomial regression after non-parametric adjustment for age and variant allele fraction as covariates. ***B.*** inset. Scatter plot with x-axis indicating the association between CH mutations and telomere lengths after adjustment for age, smoking, and genetic ancestry principal components. Association analysis with telomere lengths was performed with REGENIE.

As an orthogonal measure of clonal dynamics, we then asked how CH mutations associated with age-adjusted telomere length quantified with qPCR^38^. We reasoned that rapid clonal expansion would associate with increased attrition of telomere lengths, suggesting that the effect of CH mutations should be inversely associated with mutation fitness. Consistent with this hypothesis, we observed that age-adjusted associations with telomere lengths were negatively correlated with PACER estimates (*ρ* = −0.37, *p* = 0.016, **Figure 3B**).

Concordant with a recent report describing a telomere length attrition mechanism that is preferential to splicing factor mutations^39^, we observed that *SRSF2* and the *TERT* promoter mutations were associated with smaller reduction in telomere lengths than predicted by its fitness estimate. However, we observed that this phenomenon is broader, and likely also applies to the putative drivers in immunoglobulin loci (*IGLL5, IGH)*, underscoring the considerable heterogeneity present in clonal dynamics across the CH spectrum.

## Common disease correlates of clonal hematopoiesis

Due to an extensive literature reporting associations between CH and common aging-related disease, we then performed single-variant association analyses between CH and 30 common diseases from across the phenome. Phenotypes were defined based on “phewas” processing of the incident ICD10 codes^40^, resulting in 421,584 individuals with incident case-control information. Given the extensive possible sources of confounding in CH epidemiology^41^, we included a non-parametric specification of age at blood draw, genetic ancestry principal components, sex, and smoking history as covariates. To mitigate possible effects from cryptic relatedness, we used a generalized linear mixed model to estimate test statistics (**Methods**).

To increase power, we performed a Bayesian multi-phenotype analysis to pool signals across variants and phenotypes using mashr, an approach that has previously been shown to increase power for mapping tissue-shared QTLs^42^. Using this approach, we quantified statistical significance using Bayesian posterior estimates of the local false-sign rate (LFSR), which estimates the probability that the estimated sign of the association coefficient is correct, given the data. Overall, we performed 7,470 tests across 249 variants and 30 phenotypes (**Supplementary Data 3)**. We observed 965, 555, and 433 significant associations at LFSR thresholds of 5.0%, 1.0%, and 0.5%. At a Bonferroni threshold (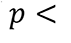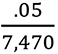), which is conservative given the dependent nature of the tests performed, we observed 72 significant mutation-phenotype associations, 92% of which (n = 66) were only observed among cancer phenotypes.

Although CH has previously been linked to several phenotypes, there are no prior reports that estimate the total variance explained on a liability scale for each phenotype – an analysis analogous to heritability estimation in complex traits. After converting effect scales to liability estimates (**Methods)**, we aggregated the variance explained by all variants for each phenotype (**Figure 4A**), which we refer to as somatic ℎ^2^estimates.

**Figure 4:**
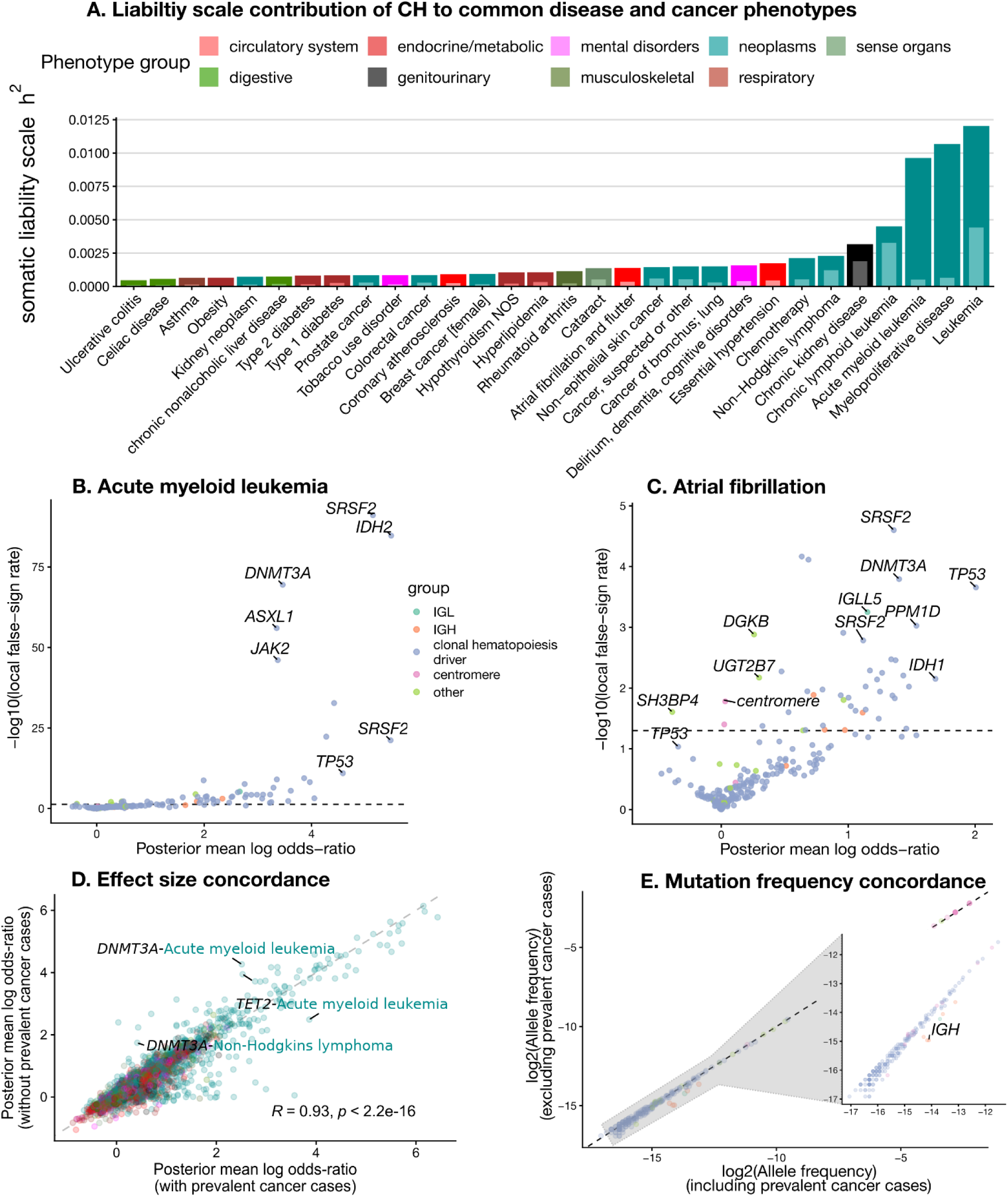
Common-disease correlates of clonal hematopoiesis (CH). **A**. Liability-scale variance explained (somatic h2) by CH across 30 incident phenotypes in UK Biobank. Bars show the summed liability-scale variance from all CH variants per phenotype, with colors denoting phenotype groups. Lighter colors denote the contribution from novel putative drivers. **B–C**. Association landscapes for acute myeloid leukemia (B) and atrial fibrillation (C). Points are CH variants colored by annotation group; the x-axis is the mashr posterior mean log odds ratio and the y-axis is −log10(local false-sign rate, LFSR). Selected loci are labeled. Positive values indicate increased risk; negative values indicate protection. The dashed horizontal line marks LFSR = 0.05. All models adjust for age at blood draw, sex, smoking, and genetic ancestry principal components, and account for relatedness via a generalized linear mixed model before multi-phenotype shrinkage with mashr. **D**. Concordance in effect sizes between PheWAS with and without individuals with prevalent cancer phenotypes. Each point is a single-variant-phenotype pair, and the x-axis indicates the posterior mean log odds-ratio from the primary PheWAS, which includes prevalent cancer cases. The y-axis indicates the posterior mean log odds-ratio when individuals with prevalent cancer (n = 38,139) were excluded. **E**. Concordance in allele frequencies between individuals with and without prevalent cancer phenotypes. Each point is a single-variant, and the x-axis indicates the allele frequency across all individuals with non-missing ICD codes (n = 421,584). The y-axis indicates the allele frequency in individuals without prevalent cancer (n = 383,445).

Concordant with previous CH phenotype analyses, we observed that the total contribution of CH was largest for hematologic malignancies (somatic ℎ^2^ = 1.2% for leukemia [phecode = 204], **Figure 4B**).

We then partitioned the variance explained between canonical CH and putative novel CH mutations (**Supplementary Data 4)**, finding that non-canonical CH mutations contributed on average 28% of the somatic ℎ^2^, ranging from 5.3% (acute myeloid leukemia) to 72.5% (chronic lymphoid leukemia). Expectedly, we found canonical CH explained the greatest variance for myeloid malignancy phenotypes, which one would expect given how canonical CH mutations were originally curated.

We observed that non-canonical CH mutations were associated with large increases in common disease risk. For example, we observed that the *UGT2B7* somatic indel was associated with increased risk for myeloproliferative neoplasms (odds-ratio of 2.1, LFSR = 1.6 x 10^-7^). Surprisingly, we also observed a substantial contribution of *IGH* mutations to atrial fibrillation risk; chr14-105863341-C-G was associated with ∼3.0 fold increase in the odds of atrial fibrillation (LFSR = 2.5 𝑥 10^−2^, **Figure 4C**). We also found that 37 of 965 significant associations (3.8%) were protective; that is, acquiring the mutation was associated with reduced risk of the disease phenotype. chr2-235126205-A-AG, an intergenic mutation ∼75kb from *SH3BP4,* was linked to 28 of these 37 protective associations. Notably, *JAK2* mutations were associated with a reduction in obesity risk (odds-ratio = 0.59, LFSR = 1.5 𝑥 10^−2^). Overall, our analysis underscores the substantial contribution of CH to hematologic and non-hematologic phenotypes, and the value of performing phenome-wide searches to place observations in context.

To interrogate the effects of including individuals with prevalent cancer (n = 38,139) in the primary PheWAS, we then performed a second PheWAS excluding all individuals with prevalent cancer, as defined by the intake surveys. We found that effect sizes were very similar (pearson 𝑟 = 0.93, **Figure 4D**, **Supplementary Data 5**), suggesting that confounding of the primary PheWAS analysis through prevalent disease or treatment at baseline is likely somewhat limited. Similarly, we found that allele frequencies were highly similar, although *IGH* mutations were modestly more prevalent in individuals that may have prevalent cancer (**Figure 4E**).

## Germline determinants of somatic *IG* mutations

Analyses of the genetic architecture of clonal hematopoiesis have revealed several common variant loci that collectively implicate HSC self-renewal, DNA repair, cell-cycle regulation, and cytokine signaling as key underlying pathways^7,17,19,21,27,30,41,43,44^. Given that previous genome-wide association studies (GWAS) have largely performed analyses that aggregate all CH mutations together (*DNMT3A / TET2 / ASLX1 / JAK2* analyses excepted), we performed a search for germline determinants of individual putative drivers. Given the substantial fitness effects of mutations in immunoglobulin loci, we first performed a GWAS of case-control phenotype for whether individuals carried at least one *IGH / IGL* mutation (n = 343 cases, 482,326 controls). After inclusion of age at blood draw, smoking history, sex, and genetic ancestry as covariates, we estimated single variant test statistics with REGENIE^45^ including both common and low-frequency (≥ 0.5%) variants. To avoid circularity from cross-mapping artifacts, we use stringent measures to exclude possible artifacts (**Methods**).

We observed a single genome-wide significant (*p* < 5 𝑥 10^−8^) locus including *GRAMD1B* (**Figure 5**). The G allele variant at this variant, rs12797056, is common (10% MAF), and was associated with greatly increased odds for an immunoglobulin mutation (odds-ratio = 1.87, *p* = 1.6 𝑥 10^−9^). We cross referenced this locus with phenome-wide association studies performed in the same cohort^46^, finding that *GRAMD1B* locus is also reported in GWAS of chronic lymphoid leukemia (CLL). Overall, these observations suggest that our variant calling workflow recapitulates a CLL linked phenotype *de novo* from sequence alone.

**Figure 5:**
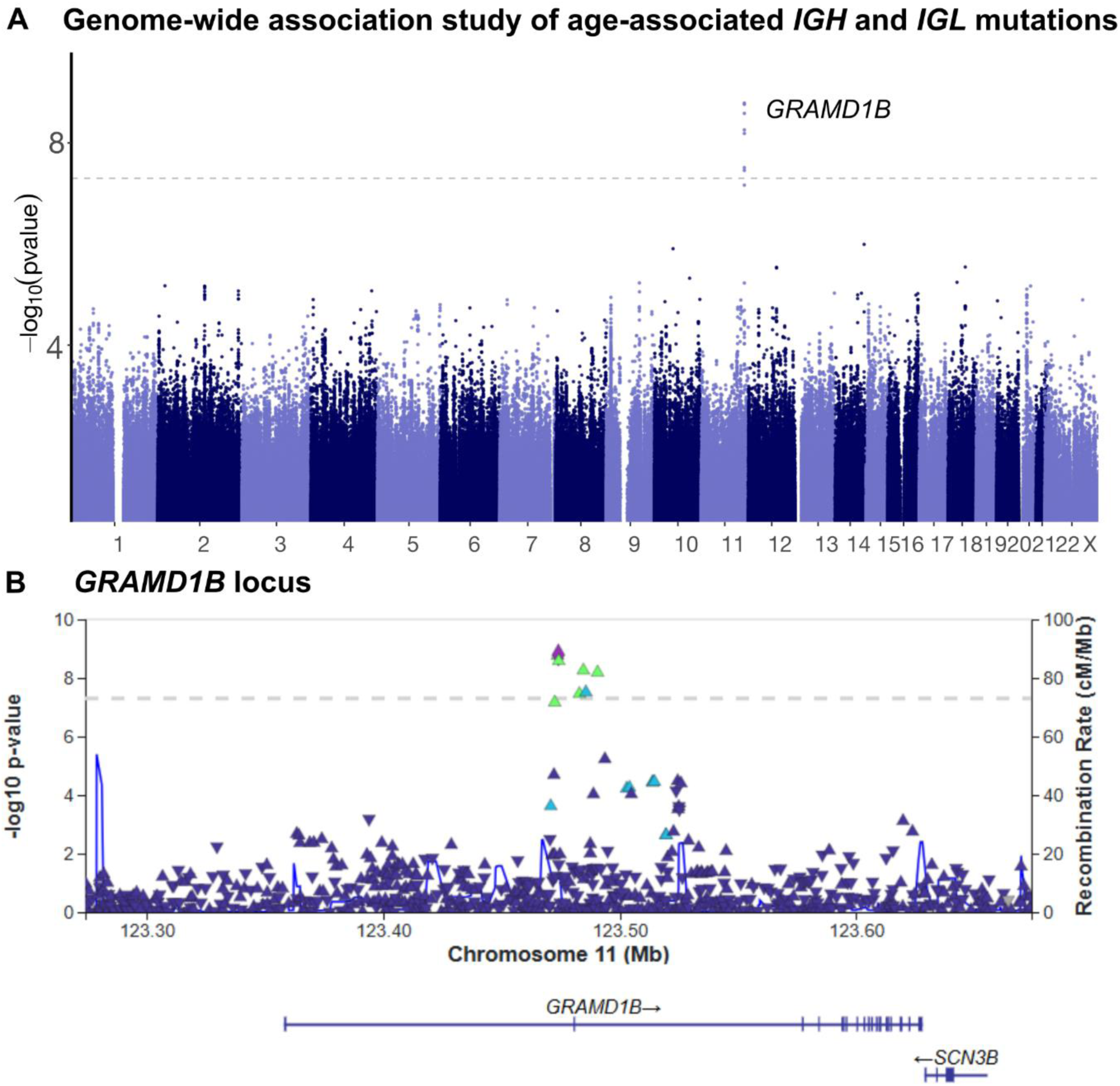
GWAS of IGH and IGL mutations (343 cases, 482,326 controls) recapitulates CLL locus. **A**. GWAS was performed with REGENIE including all variants with MAF > 0.5%. A spline of age, sex, smoking history, and genetic ancestry PCs were included as covariates. Variants falling within contigs that differed between hg19 and hg38 were excluded. Variants falling within UCSC “unusual” regions and hg38 genome reference consortium “exclusions” list were also excluded to mitigate cross-mapping artifacts. **B**. Locuszoom plot of GRAMD1B locus. LD is calculated using EUR 1000G reference samples.

We then performed a GWAS of the *UGT2B7* mutation, finding two distinct genome-wide significant loci flanking the mutation (chr4:69057335, **Supplementary Figure 8**). The first locus is in *cis* of *UGT2B7* (< 100Kb from TSS), and the lead variant (rs4348158) is low-frequency (MAF = 0.6%); the G allele is associated with greatly reduced odds of acquiring the *UGT2B7* mutation (odds-ratio = 0.26, *p* = 3.0 𝑥 10^−44^). The second locus is 800Kb downstream of the mutation, and the lead variant (rs72851490) is ∼2KB from TSS of *SULT1E1.* Large *cis* germline hits have previously been reported for *JAK2*^47,48^ and for several mCAs^17^. Both *UGT2B7* and *SULT1E* have previously been linked to sex-hormone phenotypes^45,49^; androgens have previously been shown to stimulate myelopoiesis through increased expression of *TERT* in CD34+ cells^50^. Sex-hormone binding globulin has also been nominated as a causal contributor to mosaic loss of Y in males^51^. Collectively, these observations suggest a possible sex-hormone mediated pathway through which *UGT2B7* and its germline determinants affect hematopoiesis.

## The plasma proteomic correlates of clonal hematopoiesis

Using Olink assayed proteomics measured in a subset of ∼48K individuals, we then estimated the associations between CH mutations and the circulating plasma proteome including genetic ancestry PCs, age, sex, smoking, blood cell indices, and proteomic batch as covariates (Methods). We performed this analysis on the burden of mutations within a set of selected genes (n = 14) and included 2,145 proteins for a total of 30,030 tests (14 x 2,145, **Supplementary Data 6**). To increase power, we performed a multivariate analysis with mashr, which performs a Bayesian multi-phenotype meta-analysis. We observed 897 significant associations using a Bayesian criteria (local-false sign rate < 0.05, **Figure 6**).

**Figure 6:**
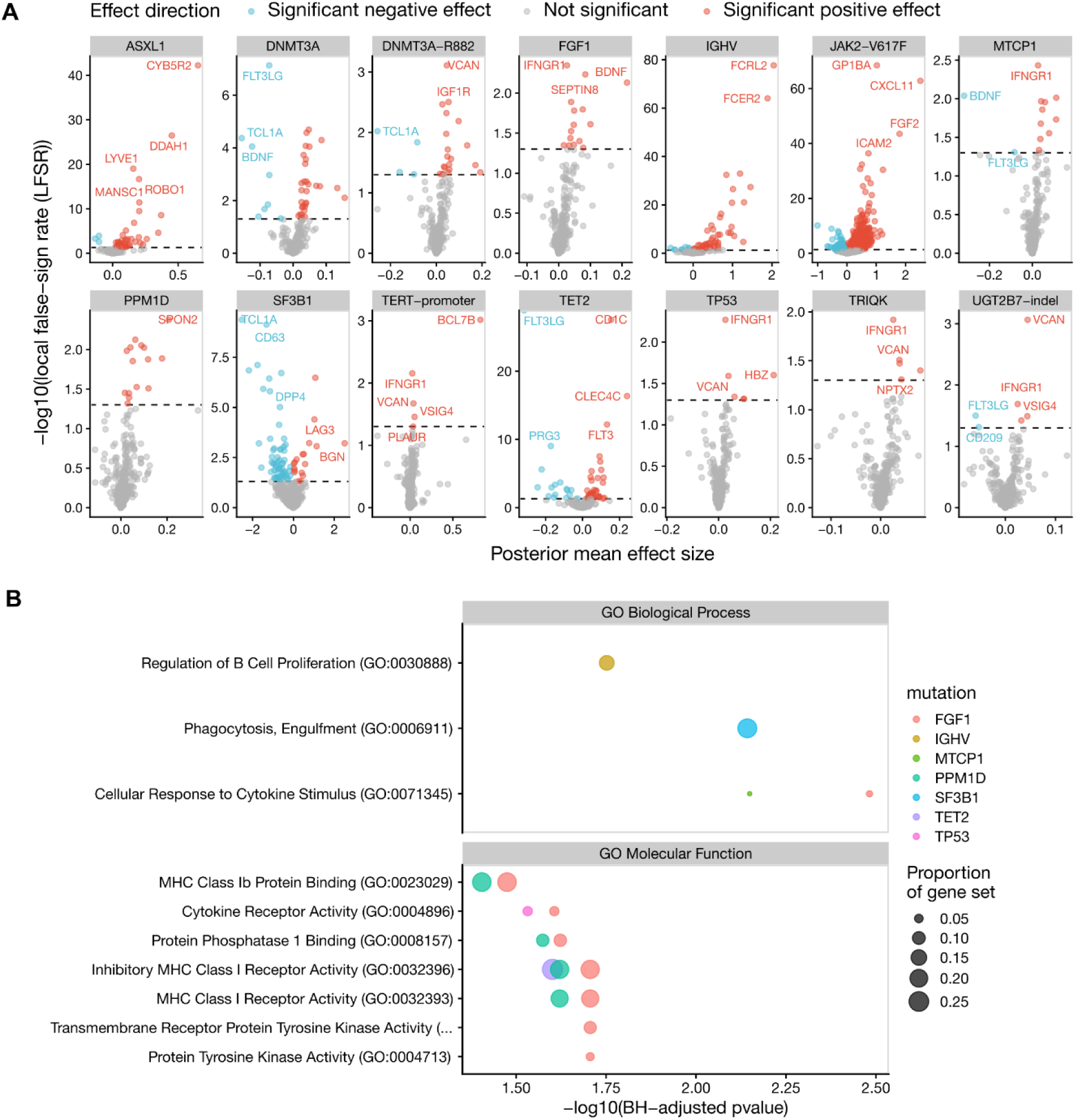
Plasma proteomic landscape of clonal hematopoiesis. **A.** Volcano plots displaying differential protein abundance associated with specific somatic mutations. The x-axis represents the posterior mean effect size, and the y-axis represents the − log_10_ local false sign rate. Each point represents a protein. The horizontal dashed line marks the significance threshold. **B.** Dot plot summarizing Gene Ontology (GO) enrichment analysis for Biological Processes (top) and Molecular Functions (bottom). The x-axis represents statistical significance and the y-axis lists the enriched GO terms. The color of the dots corresponds to the specific mutation (e.g., SF3B1 in blue, TP53 in pink), and the size of the dot represents the proportion of the gene set overlapping with the GO term.

Remarkably, 60% of these associations were due to *JAK2* (n = 541 significant hits), including the upregulation of inflammatory proteins (CXCL11, ICAM2), signaling genes (FGF2), and platelet activation (GP1BA). Although most associations were specific to a single mutation (639, or 71% of 897), two proteins were associated with all 14 mutations: IFNGR1 and VCAN. IFNGR1 is a component of the IFN𝛾 receptor. IFN𝛾 is a pro-inflammatory cytokine, and has been shown in murine models to induce excessive differentiation and to disrupt HSC quiescence^52^.

Concordant with the large effect sizes of *IGH* mutations on incident CLL risk, we observed a large association between mutated *IGH* and the abundance of B-cell specific antigens (FCER2, FCRL2, LFSR < 5.0 × 10^−60^). Concordant with a recent report examining the plasma proteomic landscape of canonical CH drivers^53^, we find that *TET2* is associated with a marked decrease in FLT3LG and an increase in FLT3; FLT3 is frequently upregulated in AML and gain-of-function FLT3 mutations are known to cause AML^54^.

Because we previously observed that *TCL1A* expression in HSCs was a potent regulator of clonal expansion^30^, we then asked whether the *TCL1A* plasma protein abundance was associated with CH. Although *TCL1A* is expressed in HSCs, it is much more highly expressed in B-cells, and thus we reasoned that *TCL1A* associations may reflect variation in B-cell clonality. Concordant with their noted roles in promoting a myeloid bias, we observed that *DNMT3A* and *SF3B1* mutations were associated with large decreases in *TCL1A* abundance, which likely reflects a decrease in B-cell counts. In contrast, *IGH* mutations were associated with large increases in plasma *TCL1A* abundance, likely reflecting a B-cell clonal expansion on which the *IGH* mutations reside.

## Discussion

Enabled by population scale whole-genome sequencing, we conducted a single-variant search for CH across the entire genome. We utilized a simple study design to search for age-associated rare variants, finding that this design was sufficient to recapitulate the majority of canonical CH driver mutations. This search revealed several non-coding point mutations beyond the *TERT* promoter, including mutations that likely affect *UGT2B7, FGF1, MTCP1/TCL1C,* and *DGKB.* We also observed clusters of age-associated mutations in *IGH* and other sites with frequent mCAs, suggesting that a scan for clustered point mutations may be sufficient to nominate mCAs. Using our genome-wide single-variant calls, we then quantified the fitness of these mutations, their germline determinants, and their common disease correlates.

To estimate the relative fitness of these mutations, we utilized an approach that we previously developed that uses passenger mutations to estimate an approximate time of mutation acquisition, which is proportional to mutation fitness in a single-mutation HSC population after adjustment for time of blood draw^30^. We observed that relative to canonical myeloid CH driver mutations, *TERT* promoter mutations had similar fitness to mutations in splicing genes and *IDH2,* which have previously been shown to be highly predictive of progression from CH to AML^55^. Stronger still were *IGH* and *IGL* mutations, which had substantially greater excess passenger mutations, suggesting that they were acquired later in life and thus expanded more rapidly to detectable clone sizes.

Concordant with recent work describing telomere attrition mechanisms for driver acquisition^39^, we observed substantial heterogeneity in the rate of telomere attrition associated with each mutation. We observed that along with *SRSF2* and *STAT3*, immunoglobulin mutations were far less likely to associate with telomere length attrition after adjustment for age at blood draw, suggesting that telomere sensitive clonal expansion mechanisms are likely more frequent than previously described. We found that the most frequent novel putative drivers were consistent with fitness estimates slightly lower than *DNMT3A-R882H^(-)^,* which would explain why they have received relatively little attention in hematologic malignancies; similar to genetic discovery in germline contexts, there is often a long-tail of non-null smaller effects sitting just below the threshold of discovery.

Although prior reports have performed PheWAS of CH^5,41,44^, none have reported an estimate of the proportion of variance explained by CH mutations on disease risk on a liability scale, which determines the possible utility of CH for population screening programs. Unsurprisingly, we find that CH contributes about an order of magnitude greater variance explained to hematologic malignancy risk than most other phenotypes. However, we also find a contribution to chronic kidney disease (CKD) that is comparable to some hematologic phenotypes, consistent with recent reports that describe links between CH and acute kidney injury^12^. In accordance with the majority of CH PheWAS reports in the UKB^5,41,44^, we find that CH explains little of the variance in coronary atherosclerosis (CAD) at a population level (<0.1% variance explained), suggesting that its population-level impact is likely multiple orders of magnitude less than traditional CAD risk factors.

Although *IGH* mutations have historically been surveilled only in lymphoid malignancies, we find that *IGH* point mutations are strongly associated with increased risk for myeloid malignancies as well, suggesting that myeloid CH analyses that exclude *IG* mutations are missing a substantial potential contributor to disease risk. Although generally considered to be restricted to B cells, our results suggest that *IGH* mutations may present frequently in myeloid progenitors as well; otherwise they would be difficult to detect with ∼30x sequencing of peripheral blood. We observed that *IGH* mutations may affect other organ systems as well, given the large increase in risk of atrial fibrillation associated with *IGH* mutations.

Intriguingly, a GWAS of a case-control *IG* phenotype defined by point mutations revealed a single significant locus at *GRAMD1B.* Given that *GRAMD1B* has previously been reported in CLL GWAS, this suggests that it may be possible to define a CLL phenotype *de novo* from sequence by examining *IG* genes for specific point mutations. As such, essentially any large-scale biobank with whole-genome sequencing is likely able to ascertain a CLL precursor phenotype.

Our study is not without limitations. First, as with any epidemiologic analysis of CH, inference of causality is challenging; in contrast to germline variation, the time at which somatic variation is acquired is generally unknown, precluding temporal ordering of disease onset and mutation acquisition. Furthermore, confounders that are known to cause CH are common and difficult to completely adjust for, including smoking effects^41^. Chronic low grade inflammation also likely has a causal effect on CH^56^, indicating likely complex causal structure implicating inflammation, CH, and several aging-related diseases. Secondly, although we report novel mutations that are age-associated – and thus indicating positive selection in HSCs -- distinguishing between bona-fide ‘drivers’ and mutations that ‘hitchhike’ on drivers remains challenging. Functional characterization of *UGT2B7, FGF1, MTCP1 / TCL1C,* and *DGKB*, which are all expressed much more highly in non-blood tissues than blood, is an important area of future work; their function here remains largely uncharacterized. Finally, although we observed extensive variation at the centromere associated with age, our sensitivity is highly limited at the centromere given the challenges of aligning short reads to highly repetitive regions. Future work using long-read sequencing in large cohorts may be required to fully characterize age associated variation at centromeres, and whether the observed centromeric point mutations alter fitness directly or merely tag centromeric instability.

Overall, we map somatic point mutations across the entire genome, finding novel disease associated mutations in parts of the genome that have typically been excluded from searches for CH driver mutations. Our summary statistics are available in a publicly available portal at somatic.emory.edu.

## Methods

### Cohorts, ethical approvals, and consent

Whole-genome sequencing (WGS) and clinical phenotypes were obtained from the UK Biobank (UKB) under application 454659. Participants provided informed consent at enrollment; the UKB study protocol received ethics approval from the UK National Research Ethics Service.

### Age association single variant analysis

A ‘gwas’ of age at blood draw was performed using all variants present in “/Bulk/DRAGEN WGS/DRAGEN population level WGS variants, PLINK format [500k release]/” plink files. All variants with at least 10 minor alleles were included, and sex, ‘ever smoker’, and the first 10 genetic ancestry PCs were included as covariates. Regenie^45^ was used to perform association analyses. Age associated variants were declared at a genome-wide significance threshold.

### Gene-based association analyses

Two kinds of gene-based analyses were performed. First, single-variant pvalues were meta-analyzed within all protein-coding genes with ACAT^35^, including all variants with minor allele frequencies < 0.5% were included. Secondly, an RVAS was performed by using only rare (< 0.5% MAF) variants with AlphaMissense pathogenicity scores greater than 0.3.

Pvalues were again meta-analyzed with ACAT.

### Loss of Y calling

Mosaic loss of Y was called on the whole-genomes by first quantifying sequencing depth at a 12 pre-specified regions using Mosdepth^57^, including three on the X chromosome, three on the Y chromosome, the *DNMT3A* locus, *TET2* locus, *JAK2* locus, *OCT4* locus, *ASXL1* locus, and the chr17 centromere. Depth was then normalized per sample by dividing by the depth at the *OCT4* locus using the following WDL: https://github.com/weinstockj/mca_calling_workflow. Mosaic loss of Y was defined in males as samples with a relative Y chromosome depth of less than 0.369.

### Variant calling of canonical CH mutations

Non-reference alleles were called at a set of pre-specified list of 4,737 variants defined based on the union of CH mutations detected in TOPMed^43^ and a previous UKB analysis^5^. Variants were then called on the UKB exomes at these sites using a bcftools mpileup approach:

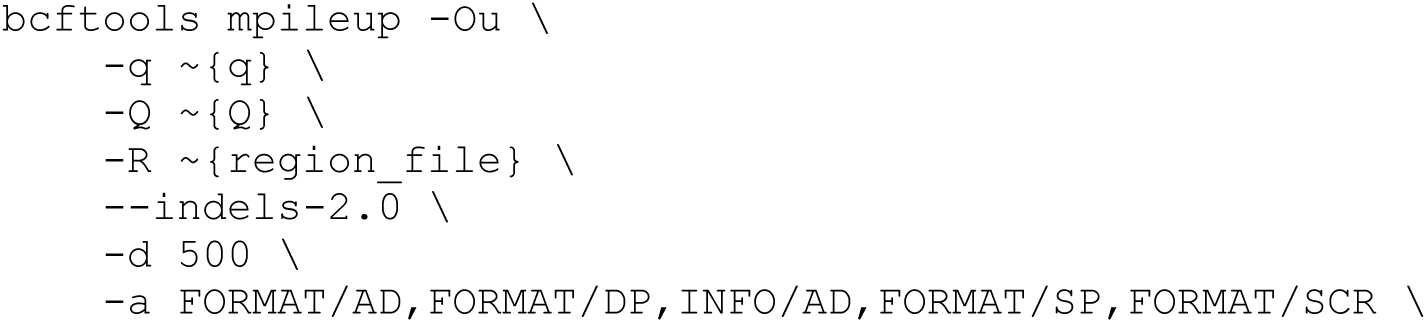
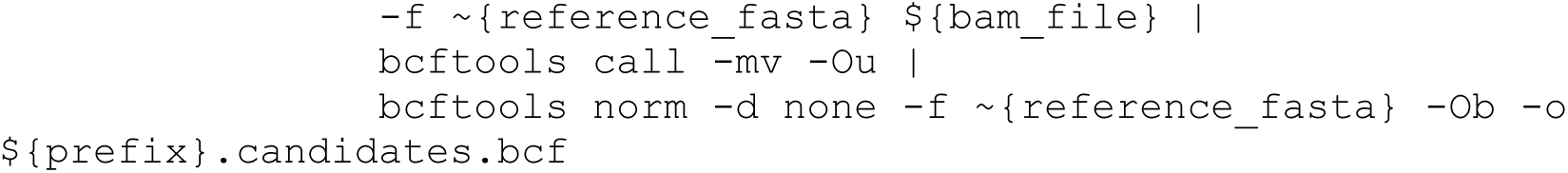

Where “q”, the minimum mapping quality of the alignment was set to 10, and “Q”, the minimum base quality was set to 20. After preliminary calling, all variants with mean variant allele fractions (VAFs) between [.46, .54] were excluded, which totaled 577 variants. Canonical mutations were detected in 18,010 individuals.

### Variant calling of putative novel drivers

After identifying age-associated variants, we then extracted variant allele fractions and other QC metrics from the Dragen single sample VCFs at the specified 45 variants, using a custom Rust binary (https://github.com/weinstockj/parseSomaticDragen).

### Variant allele fraction hypothesis testing

To assess whether the observed variant allele fraction deviates from the germline heterozygous expectation, we conducted a beta–binomial likelihood ratio test (LRT). The beta–binomial model accommodates extra-binomial variation in read counts.

Under the null hypothesis (*p*_0_ = 0.5) and the alternative (*p*_1_ = vaf), log-likelihoods were computed using the beta–binomial density with overdispersion parameter *ρ* = 0.1(VGAM package):

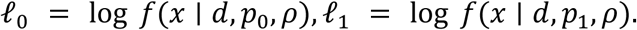

Depth (𝑑) was fixed at 30x. To aggregate evidence across the cohort for each variant, the log-likelihood difference was scaled by the observed alternate-allele count 𝑛:

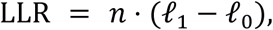

and the LRT statistic was formed as:

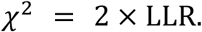

P-values were obtained from a chi-squared distribution with 1 degree of freedom.

### Passenger burden quantification

Passengers were quantified by parsing the Illumina Dragen UKB pVCFs “/Bulk/DRAGEN WGS/DRAGEN population level WGS variants, pVCF format [500k release]”. pVCFs were first filtered to bi-allelic singleton SNPs (mac = 1). Then, using a custom Rust binary (https://github.com/weinstockj/count_somatic), output was filtered to variants with variant allele fractions between [0.05, 0.35], and finally again filtered to variants with the number of alt reads between [4, 6].

### PACER gene level estimates

Relative fitness was estimated using PACER, which leverages enrichment of hitchhiking passenger mutations to infer the relative time of driver acquisition and clone growth.

Briefly, excess passengers per clone were modeled with negative binomial regression, adjusting for age at blood draw and observed VAF with natural splines.

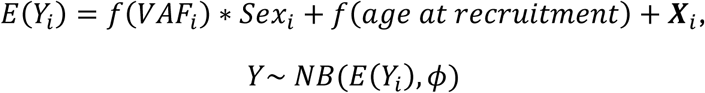

Where *f*(*x*) denotes a natural spline, and 𝑿_𝒊_ denotes an indicator function for mutation carrier status.

PACER estimates are reported relative to *DNMT3A* R882H (chr2:25234373 C>T).

### Telomere length estimates

Regenie was used to compute test statistics for an association analysis between CH mutations and qPCR quantified telomere lengths, which have been previously adjusted for technical covariates^38^ (“z_adjusted_telomere_ratio”). A natural spline of age, sex, “ever smoker”, and the first 10 genetic ancestry PCs were included as covariates.

### Phenome-wide association study

ICD10 codes were extracted and processed with the “phecode” PheWAS mapping^40^. 30 common diseases were selected for analysis. Single-variant association analyses were performed using CH point mutations with at least 10 minor alleles using Regenie. Smoking history, a spline of age, sex, and the first 10 genetic ancestry PCs were included as covariates. After obtaining single-phenotype analyses, a multivariate analysis was performed with mashr^42^, which also estimates posterior estimates of log odds-ratios, qvalues, and local false-sign rates^58^.

We then performed a second PheWAS after excluding individuals with prevalent cancer at baseline. Prevalent cancer (n = 38,139 individuals) was defined from self-reported health information obtained at enrollment.

### Liability scale variance explained

To quantify the contribution of clonal hematopoiesis (CH) mutations to disease risk, we first estimated the observed variance of each phenotype on the binary scale. For each phenotype, the prevalence (*p*) was estimated based on the prevalence in the UKB cohort.

Next, we converted single-variant effect sizes from odds ratios (OR) to liability-scale beta coefficients using a threshold model under a probit approximation. For each variant, the liability-scale effect 𝛽_𝑙𝑖𝑎𝑏𝑖𝑙𝑖𝑡𝑦_was derived from a log odds ratio conversion to probit scale.

We aggregated the liability-scale variance explained across all CH variants for each phenotype using:

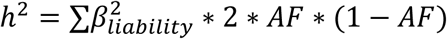

This calculation was performed separately for (i) all CH variants and (ii) novel non-canonical CH variants (excluding known CHIP genes). Covariance between mutations was ignored as an approximation.

### GWAS

GWAS of the *IG* and *UGT2B7* mutation phenotypes were performed with Regenie. A spline of age, sex, smoking history, and the first 10 genetic ancestry PCs were included as covariates. All variants were included with minor allele frequencies of at least 0.5%.

Variants were then restricted to those were marked “PASS” among variants with at least 10 minor allele counts in TOPMed Freeze 12^33^. Three additional exclusion lists were then

applied:

1. Variants falling within contigs that differed between hg19 and hg38
2. Variants in “UCSC unusual” regions
3. Variants in the GRC exclusions list for hg38

### Protein-wide association study

For each mutation, we tested for association between carrier status and circulating plasma protein abundance using ordinary least squares linear regression across assayed proteins, retaining the 75% of proteins with the most non-missing observations. Covariates included sex, age at recruitment, smoking status (ever smoked), the first 10 genetic principal components (gPC1–gPC10), complete blood count variables (monocyte, neutrophil, lymphocyte, eosinophil and platelet counts; and proteomics technical variables (number of proteins measured, well identifier used for the sample run, and volume of serum held). Age at recruitment was modeled using a natural cubic spline basis.

To obtain Bayesian shrinkage estimates and false sign rate control we applied multivariate adaptive shrinkage (mashr). We constructed a matrix of effect estimates (proteins × mutations) and a corresponding matrix of standard errors. We fit mashr using the canonical covariance matrices to capture patterns of sharing across mutations and extracted mutation-protein local false-sign rates (LFSR), posterior mean and posterior standard deviation from the fitted model.

## Code availability

1. Regenie WDL workflow: https://github.com/weinstockj/UKB_regenie_workflow
2. Illumina Dragen VCF to parquet: https://github.com/weinstockj/parseSomaticDragen
3. Loss of Y calling: https://github.com/weinstockj/mca_calling_workflow
4. CHIP calling: https://github.com/weinstockj/UKB_CHIP_calling
5. Variant annotation, filtering, plotting: https://github.com/weinstockj/gwasplot
6. Passenger mutation Rust binary: https://github.com/weinstockj/count_somatic
7. Passenger mutation calling WDL: https://github.com/weinstockj/count_somatic_workflow

## Data Availability

UKB data were accessed under application number 454659. Summary statistics for PheWAS analyses are available at a public web portal, somatic.emory.edu. GWAS summary statistics for the *IG* GWAS are available here: https://my.locuszoom.org/gwas/493597/.

## Data Availability

UKB data is accessible via application through https://ams.ukbiobank.ac.uk/ams/. Summary statistics for PheWAS analyses are available at a public web portal, somatic.emory.edu. GWAS summary statistics for the IG GWAS are available here: https://my.locuszoom.org/gwas/493597/.

https://somatic.emory.edu/

## Acknowledgements

UKB data were accessed under application number 454659. JSW acknowledges startup support from the Emory University School of Medicine and the Winship Cancer Institute. The authors also thank Po-Ru Loh for insightful conversations on analyses.

## Supplementary Figures

**Supplementary Figure 1:**
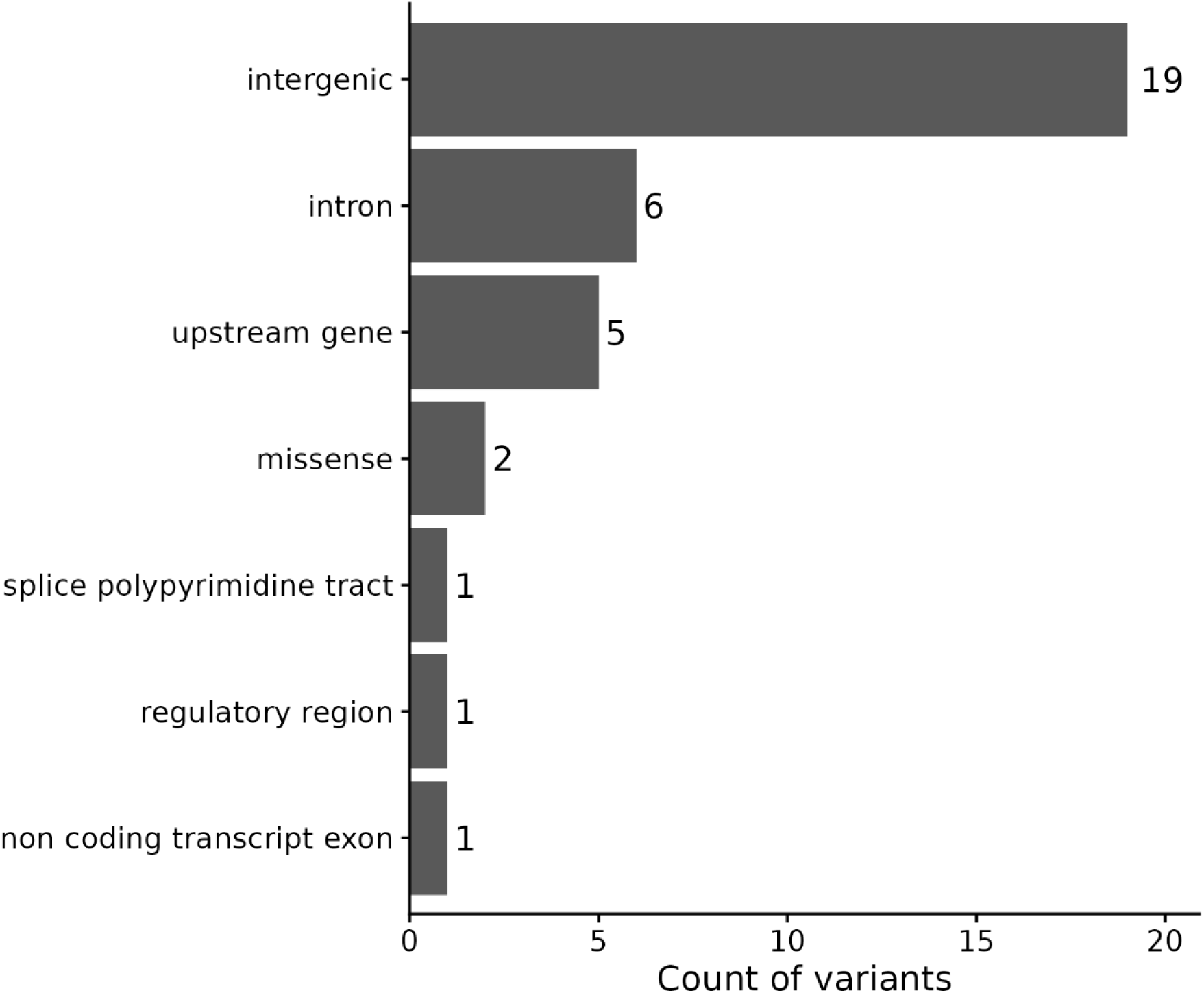
Distribution of functional annotations for 35 novel age-associated variants, as annotated by the Ensembl variant effect predictor. Bars indicate counts of missense, splicing, and non-coding variants.

**Supplementary Figure 2:**
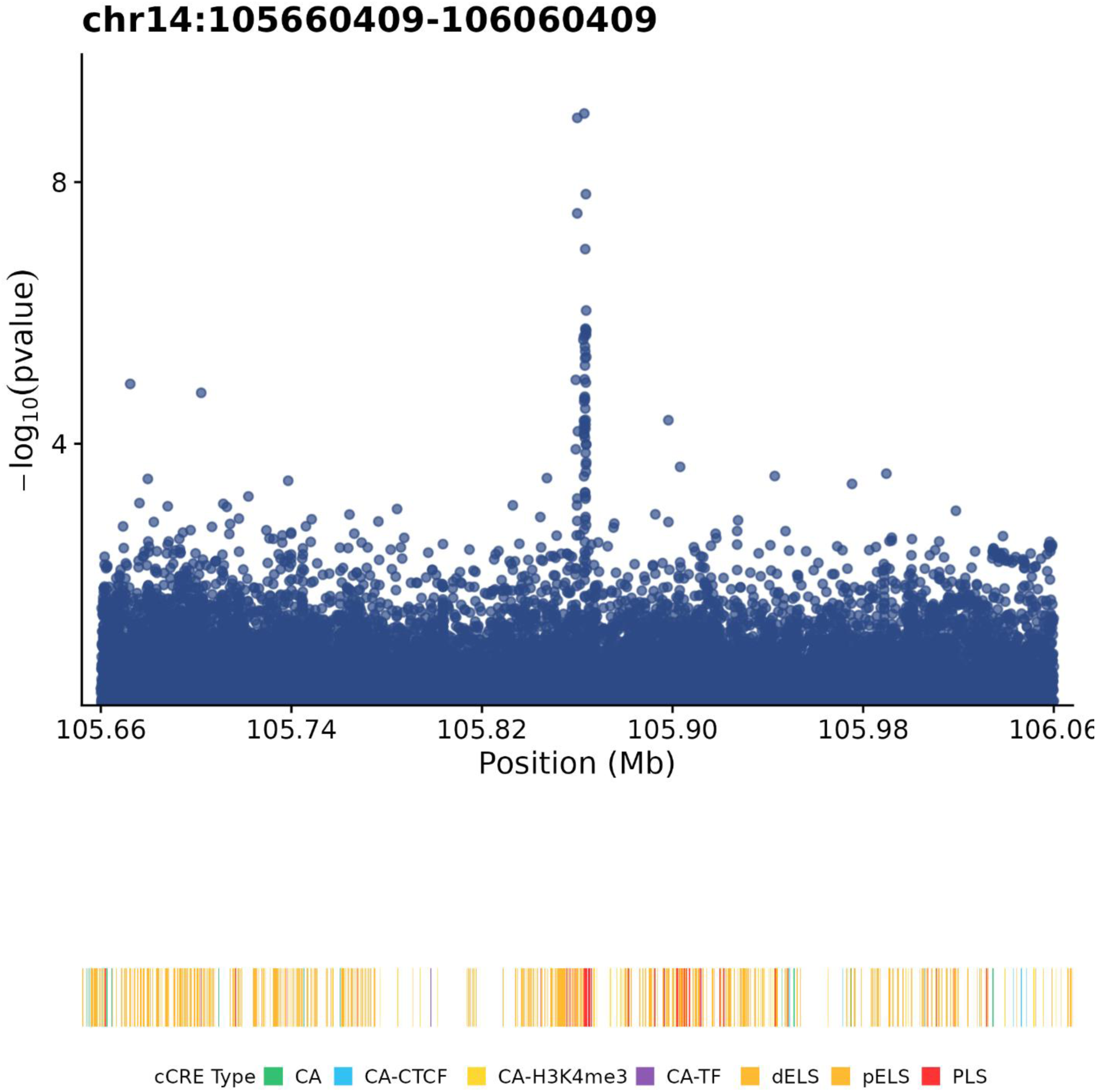
View of IGH locus showing five age-associated point mutations. Track on the bottom displays cCRE elements from ENCODE for hematopoietic multipotent progenitor biosample ENCDO116AAA.

**Supplementary Figure 3:**
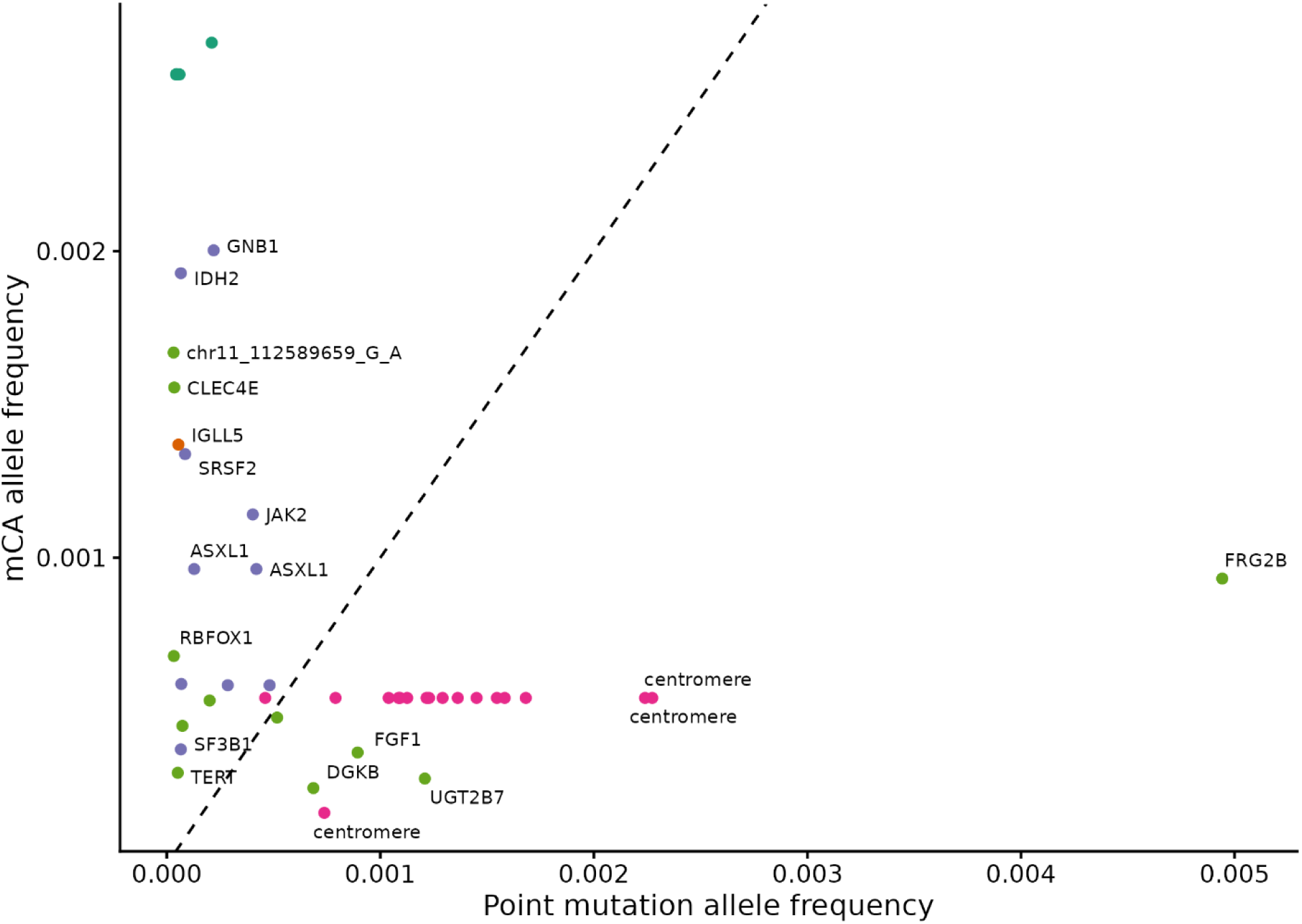
X-axis indicates the frequency of point mutations in UKB. Y axis indicates the frequency of overlapping mCAs as reported in Loh et al., 2020.

**Supplementary Figure 4:**
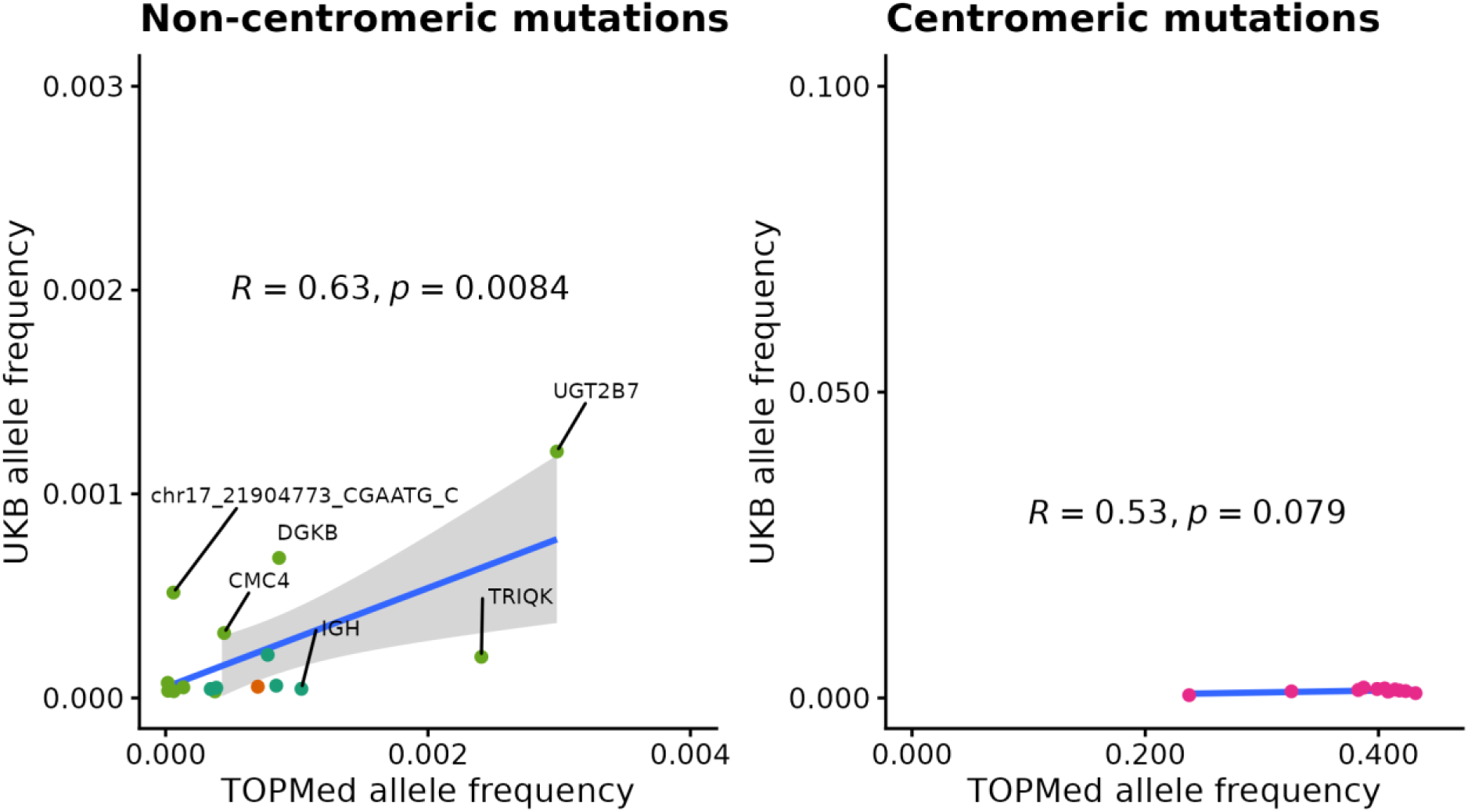
Scatter plot of alternative allele frequencies for 45 age-associated variants across UK Biobank and TOPMed cohorts. Non-centromeric variants show strong concordance (ρ = 0.63, p = 0.008), while centromeric variants exhibit higher frequencies in TOPMed.

**Supplementary Figure 5:**
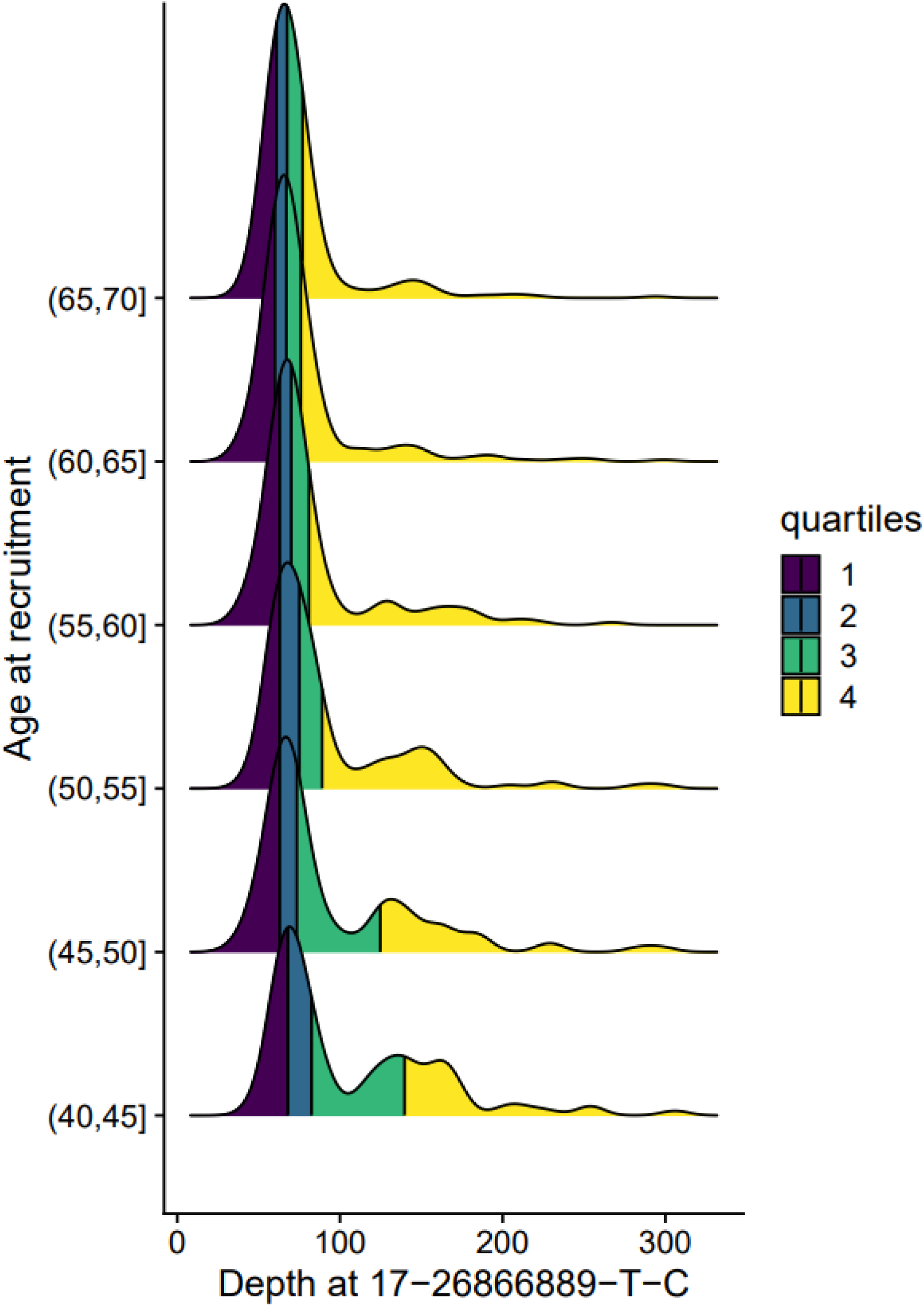
Y-axis indicates the density of age at blood draw; colors indicate quartiles of the distribution. X-axis indicates the sample level sequencing depth at 17-26866889-T-C.

**Supplementary Figure 6:**
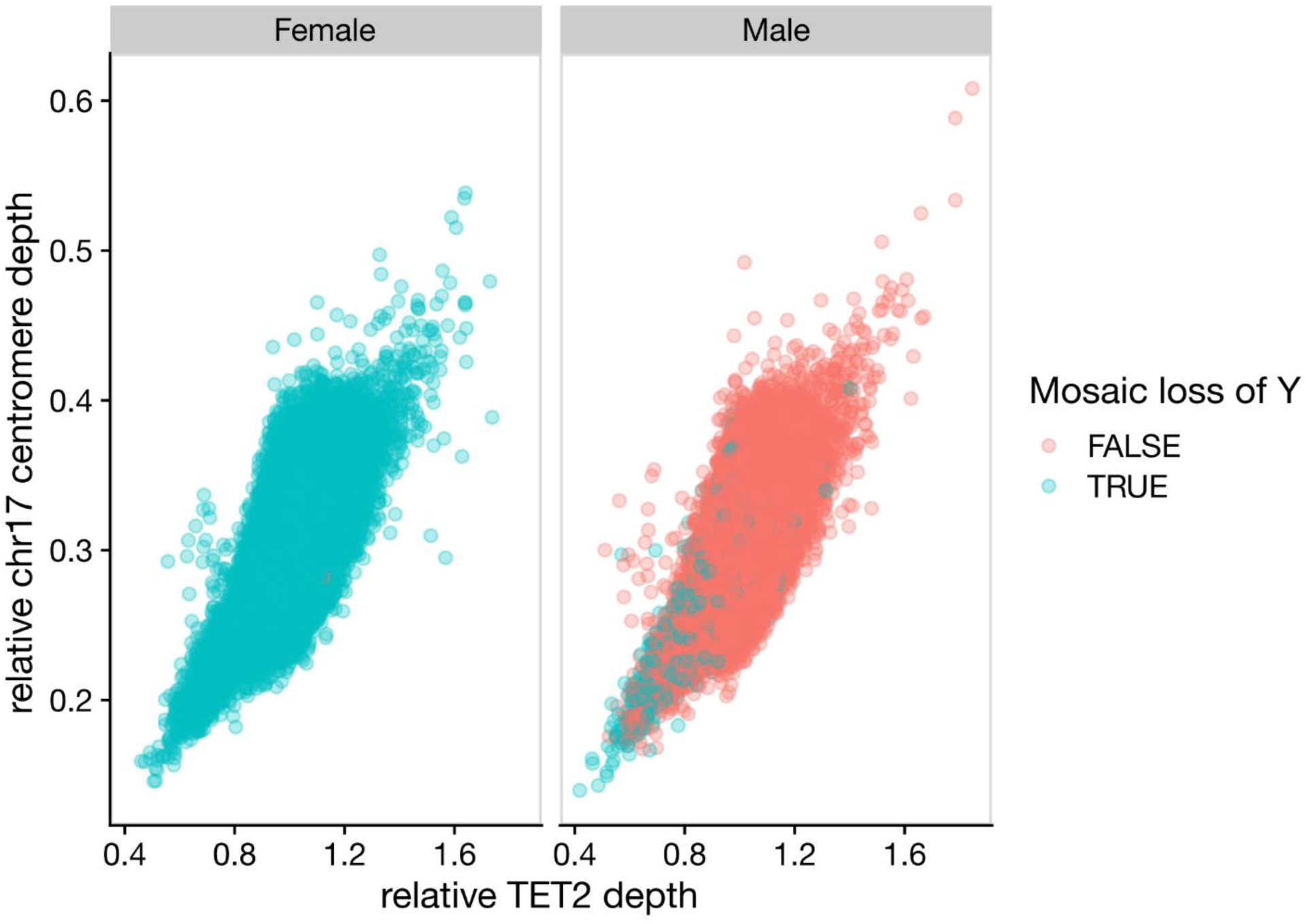
Depth was quantified using Mosdepth at the chr 17 centromere, OC4, TET2, and sex chromosomes. Depths were then “normalized” by dividing by the depth at OCT4. Mosaic loss of Y was called in males by thresholding the relative Y chromosome depth below 0.369.

**Supplementary Figure 7:**
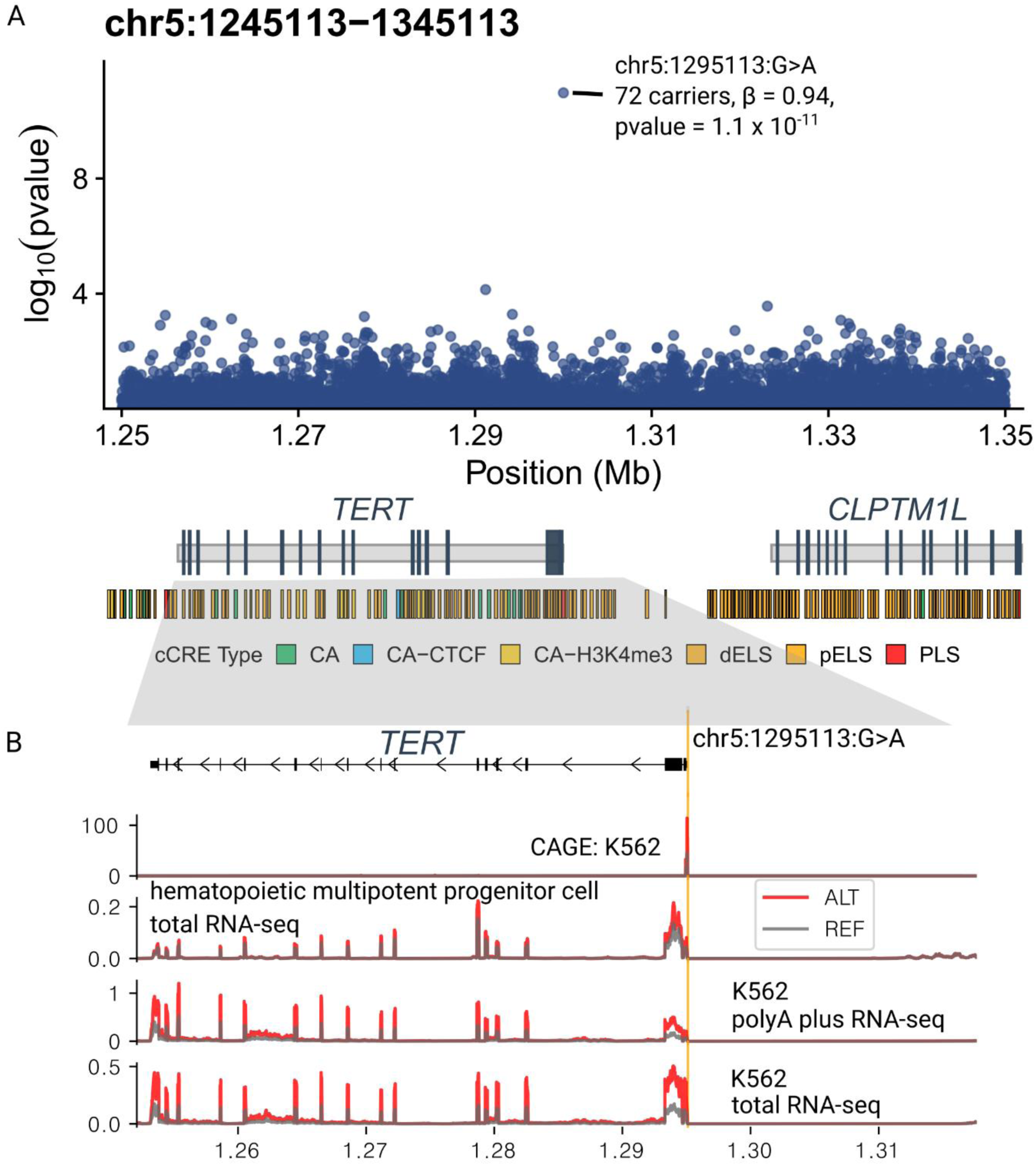
A. Locuszoom of association between age at blood draw and variants (both germline + somatic) in the TERT locus. The age-associated putative driver, chr5:1295113:G>A is highlighted. B., AlphaGenome predicted RNA-seq tracks in the locus.

**Supplementary Figure 8:**
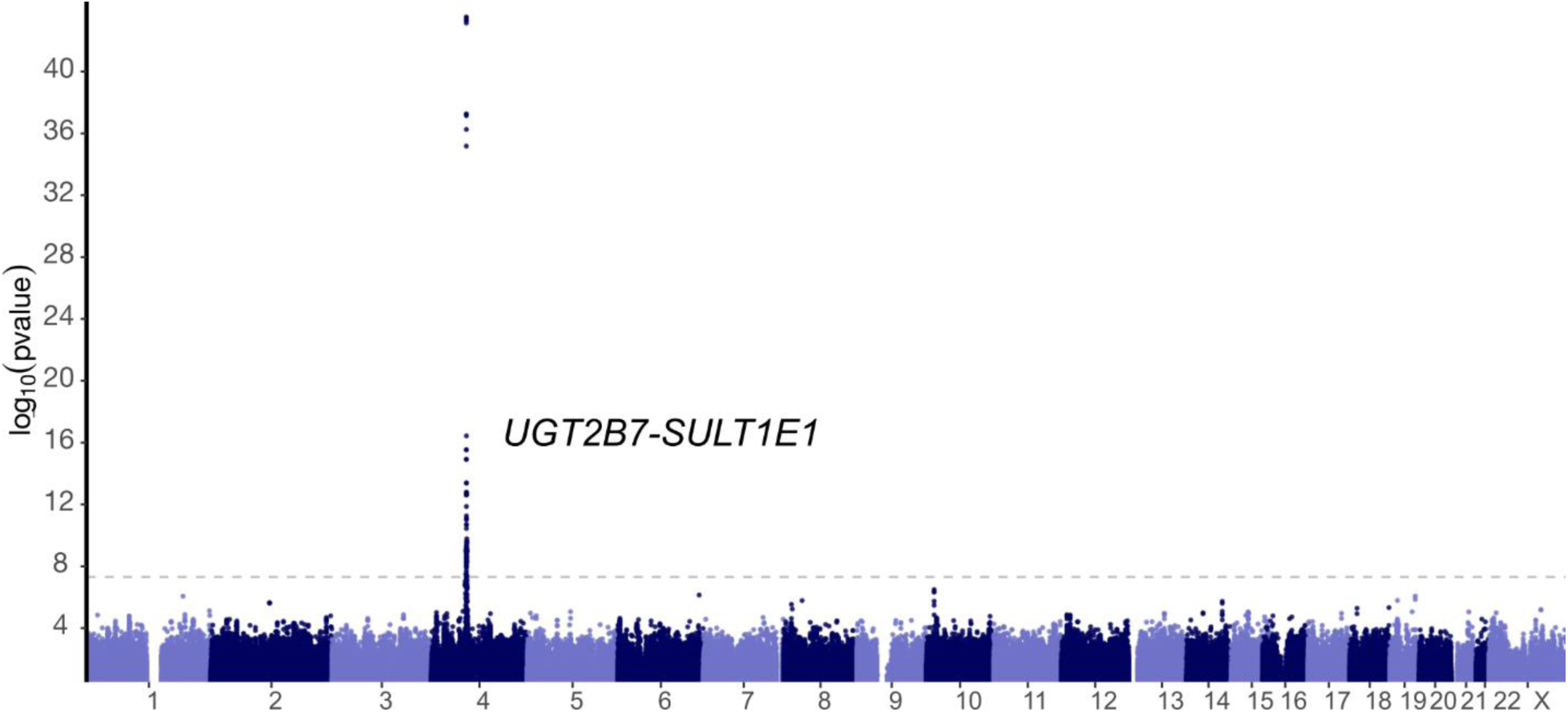
GWAS was performed with REGENIE including all variants with MAF > 0.5%. A spline of age, sex, smoking history, and genetic ancestry PCs were included as covariates. Variants falling within contigs that differed between hg19 and hg38 were excluded. Variants falling within UCSC “unusual” regions and hg38 genome reference consortium “exclusions” list were also excluded to mitigate cross-mapping artifacts.

